# Early Anakinra Treatment for COVID-19 Guided by Urokinase Plasminogen Receptor

**DOI:** 10.1101/2021.05.16.21257283

**Authors:** Evdoxia Kyriazopoulou, Garyfallia Poulakou, Haralampos Milionis, Simeon Metallidis, Georgios Adamis, Konstantinos Tsiakos, Archontoula Fragkou, Aggeliki Rapti, Christina Damoulari, Massimo Fantoni, Ioannis Kalomenidis, Georgios Chrysos, Andrea Angheben, Ilias Kainis, Zoi Alexiou, Francesco Castelli, Francesco Saverio Serino, Petros Bakakos, Emanuele Nicastri, Vassiliki Tzavara, Evangelos Kostis, Lorenzo Dagna, Panagiotis Koufargyris, Katerina Dimakou, Glykeria Tzatzagou, Maria Chini, Matteo Bassetti, Konstantina Katrini, Vasileios Kotsis, George Tsoukalas, Carlo Selmi, Ioannis Bliziotis, Michael Samarkos, Michael Doumas, Sofia Ktena, Aikaterini Masgala, Ilias Papanikolaou, Aikaterini Argyraki, Chiara Simona Cardellino, Eleni-Ioanna Katsigianni, Efthymia Giannitsioti, Antonella Cingolani, Karolina Akinosoglou, Orestis Liatsis-Douvitsas, Styliani Symbardi, Maria Mouktaroudi, Giuseppe Ippolito, Eleni Florou, Antigone Kotsaki, Mihai G. Netea, Jesper Eugen-Olsen, Miltiades Kyprianou, Periklis Panagopoulos, George N. Dalekos, Evangelos J. Giamarellos-Bourboulis

## Abstract

**Background:** In a previous open-label trial, early anakinra treatment guided by elevated soluble urokinase plasminogen activator receptor (suPAR) prevented progression of COVID-19 pneumonia into respiratory failure.

**Methods:** In the SAVE-MORE multicenter trial, 594 hospitalized patients with moderate and severe COVID-19 pneumonia and plasma suPAR 6 ng/ml or more and receiving standard-of-care were 1:2 randomized to subcutaneous treatment with placebo or 100 mg anakinra once daily for 10 days. The primary endpoint was the overall clinical status of the 11-point World Health Organization ordinal Clinical Progression Scale (WHO-CPS) at day 28. The changes of the WHO-CPS and of the sequential organ failure assessment (SOFA) score were the main secondary endpoints.

**Results:** Anakinra-treated patients were distributed to lower strata of WHO-CPS by day 28 (adjusted odds ratio-OR 0.36; 95%CI 0.26-0.50; P<0.001); anakinra protected from severe disease or death (6 or more points of WHO-CPS) (OR: 0.46; P: 0.010). The median absolute decrease of WHO-CPS in the placebo and anakinra groups from baseline was 3 and 4 points respectively at day 28 (OR 0.40; P<0.0001); and 2 and 3 points at day 14 (OR 0.63; P: 0.003); the absolute decrease of SOFA score was 0 and 1 points (OR 0.63; P: 0.004). 28-day mortality decreased (hazard ratio: 0.45; P: 0.045). Hospital stay was shorter.

**Conclusions:** Early start of anakinra treatment guided by suPAR provides 2.78 times better improvement of overall clinical status in moderate and severe COVID-19 pneumonia.

(Sponsored by the Hellenic Institute for the Study of Sepsis ClinicalTrials.gov identifier, NCT04680949)

## INTRODUCTION

COronaVIrus Disease-19 (COVID-19) pneumonia can have an unpredictable clinical course. Patients may suddenly deteriorate into severe respiratory failure (SRF), so that early recognition of the turning point and timely onset of targeted treatment appear of outmost importance.

Our group and others have shown that soluble urokinase plasminogen activator (suPAR) can early prognosticate unfavorable outcome^1, 2^. Early suPAR increase is an indicator of the release of danger-associated molecular patterns (DAMPs), namely calprotectin (S100A8/A9) and interleukin (IL)-1 α^3,4^. Calprotectin in turn stimulates the aberrant production of interleukin (IL)-1β by the circulating monocytes^4^ whereas knock-outing of IL-1α is protective for the host^3^. These observations frame the hypothesis that early detection of increased suPAR may guide targeted therapeutics against IL-1α and IL-1β. Indeed, in the open-label phase II study SAVE, early administration of anakinra guided by suPAR decreased the relative risk for progression into SRF by 70% compared to standard-of-care treatment; significant reduction of 28-day mortality was also found. The recombinant IL-1 receptor antagonist anakinra blocks both IL-1α and IL-1β^5^.

Despite the important information provided by the SAVE trial, one prospective randomized clinical trial (RCT) is necessary to prove the effectiveness of this approach. SAVE-MORE (<u>s</u>uPAR-guided <u>A</u>nakinra treatment for <u>V</u>alidation of the risk and <u>E</u>arly <u>M</u>anagement <u>O</u>f seve<u>RE</u> respiratory failure by COVID-19) is a pivotal, confirmatory, phase III RCT aiming to evaluate the efficacy and safety of early start of anakinra guided by suPAR in patients with COVID-19 pneumonia. The primary objective was to evaluate the efficacy and safety of early targeting of IL-1α/β on the clinical state of patients with COVID-19 pneumonia and elevated suPAR levels, over 28 days, as measured by the 11-point ordinal WHO (World Health Organization) Clinical Progression scale (CPS).

## METHODS

### Trial oversight

SAVE-MORE is a prospective double-blind RCT conducted in 37 study sites (29 in Greece and 8 in Italy). The protocol (available with the full text of this article) was finalized after advice by the Emergency Task Force (ETF) of the European Medicines Agency (EMA) for COVID-19 (document EMA/659928/2020). The protocol was approved by the National Ethics Committee of Greece (approval 161/20) and by the Ethics Committee of the National Institute for Infectious Diseases Lazzaro Spallanzani, IRCCS in Rome (01.02.2021) (EudraCT number, 2020-005828-11; ClinicalTrials.gov NCT04680949). The trial was sponsored by the Hellenic Institute for the Study of Sepsis (HISS) and funded in part by HISS and in part by Swedish Orphan Biovitrum AB (Sobi). HISS was responsible for the design, conduct, analysis and interpretation of data, and decision to publish. The laboratory of Immunology of Infectious Diseases of the 4^th^ Department of Internal Medicine at ATTIKON University General Hospital served as central laboratory. The data lock for all events until day 28 was done on April 29^th^ 2021.

### Patients

Enrolled patients were adults of either gender; with molecular diagnosis of infection by SARS-CoV-2; with involvement of the lower respiratory tract as confirmed by chest computed tomography or X-ray; in need for hospitalization; and with plasma suPAR 6 ng/ml or more. Main exclusion criteria were: ratio or partial oxygen pressure to fraction of inspired oxygen less than 150; need of non-invasive ventilation (CPAP or BPAP) or mechanical ventilation; neutropenia; stage IV malignancy; end-stage renal disease; severe hepatic failure; immunodeficiencies; and chronic intake of corticosteroids and biological anti-cytokine drugs. All patients or their legal representatives provided written informed consent before enrollment.

### Trial interventions

Patients meeting all inclusion criteria and not meeting any exclusion criterion were subject to blood draw. suPAR was measured in plasma using the suPARnostic^®^ Quick Triage kit (Virogates S/A, Birkerød, Denmark) and a point-of care reader. Patients with suPAR 6 ng/ml or more were electronically 1:2 randomized into treatment with placebo or anakinra using four randomization strata: classification into moderate or severe disease using the WHO definition^6^; need for dexamethasone intake; body mass index (BMI) more than 30 kg/m^2^; and country. The study drug was administered subcutaneously once daily in the thigh or in the abdomen for seven to 10 days. Patients allocated to placebo treatment were daily injected 0.67 ml of 0.9% sodium chloride; and those allocated to active drug 100 mg of anakinra at a final volume of 0.67 ml. Study drug was prepared by an unblinded pharmacist with access to the electronic study system using a separate username and a password. Administration was done by a blind study nurse. All patients were receiving pre-defined standard-of-care (SoC) which consisted of regular monitoring of physical signs, oximetry and anti-coagulation. Patients with severe disease by the WHO definition^6^ were also receiving intravenous 6 mg daily dexamethasone for 10 days. Remdesivir treatment was left at the discretion of the attending physicians; other biologicals targeting cytokines and kinase inhibitors were not allowed.

Study visits were done daily for 10 days; on day 14; and on day 28. At each study-visit the following were recorded: non-serious and serious treatment-emergent adverse events (TEAEs); WHO-CPS; sequential organ failure assessment (SOFA) score; and co-administered treatment. Visits were done by phone for patients discharged by day 7. Data were captured after review of all medical and nursing charts by a physicians’ team blinded to the allocation group. Blood samples and nasopharyngeal swabs were collected before start of the study drug and at days 4 and 7 for the measurements of biomarkers.

All serious and non-serious TEAEs were graded according to the Common Terminology Criteria for Adverse Events (version 5.0).

### Outcomes

The primary study endpoint was the overall comparison of the distribution of frequencies of the scores from the 11-point WHO Clinical Progression ordinal Scale (CPS) between the two arms of treatment at Day 28. Secondary endpoints included the changes of WHO-CPS by days 14 and 28 from the baseline (before start of the study drug); the change of SOFA score by day 7 from baseline; the time until hospital discharge; the time of stay in the intensive care unit (ICU) for patients eventually admitted to the ICU; and the comparison of biomarkers.

### Statistical analysis

The sample size was calculated based on the finding from the phase II SAVE trial^5^ that 42% of comparators and 16.3% of anakinra-treated patients by day 28 were presented with 6 or more points of the WHO-CPS. To achieve such a difference in the WHO-CPS scores with 90% power at the 5% level of significance, allocation of 200 patients to SoC and placebo treatment and 400 patients to SoC and anakinra treatment were planned. Data were analyzed for the intention-to-treat (ITT) population. Missing data were imputed by last observation carried forward (LOCF). WHO-CPS is an ordinal 11-point variable ranging from 0 to 10 and comparisons were done by univariate and multivariate ordinal regression analysis using logit function. Results were expressed as the odds ratio (OR) and 95% confidence intervals (CI). The two basic assumptions of the model, i.e., proportional odds and the goodness-of-fit test were checked. According to EMA’s COVID-ETF advice the variables used for stratified randomization entered as co-variates in the multivariate model, i.e., disease severity, intake of dexamethasone, BMI more than 30, and country. According to the same advice, the analysis of the primary endpoint should have been supported by three analyses: comparison of the WHO-CPS by day 14; logistic regression analysis separately for patients at the two spectra of WHO-CPS at day 28; and time progression to respiratory failure by day 14. The first spectrum of the WHO-CPS was defined as patients fully recovered with negative viral load (WHO-CPS 0 points) contrary to patients with persistent disease (WHO-CPS between points 1 to 10). The second spectrum was defined as patients pointed 6 or more in the WHO-CPS (severe hospitalized and dead) contrary to patients pointed 5 or less. Five sensitivity analyses were conducted to assess robustness: exclusion of population deviating from the SoC; population receiving at least 7 doses of the study drug; complete analysis set; responder analysis treating missing values as non-responders; and comparison of the unadjusted and the adjusted treatment effects. Analysis was conducted using IBM SPSS Statistics v. 26.0. All P values were two-sided and any P value <0.05 was considered as statistically significant. The complete statistical analysis plan is provided in the Supplementary Appendix.

## RESULTS

### Patients

From December 2020 through March 2021, 1060 patients were screened and 606 were randomized. 12 patients withdrew consent and requested removal of all data, leaving a final ITT analysis cohort of 594 patients; 189 patients were allocated to the SoC and placebo arm, and 405 patients were allocated to the SoC and anakinra arm. Only one patient was lost to follow-up (Figure 1). Baseline characteristics and co-administered treatments were similar between the two arms (Table 1).

**Figure 1.**
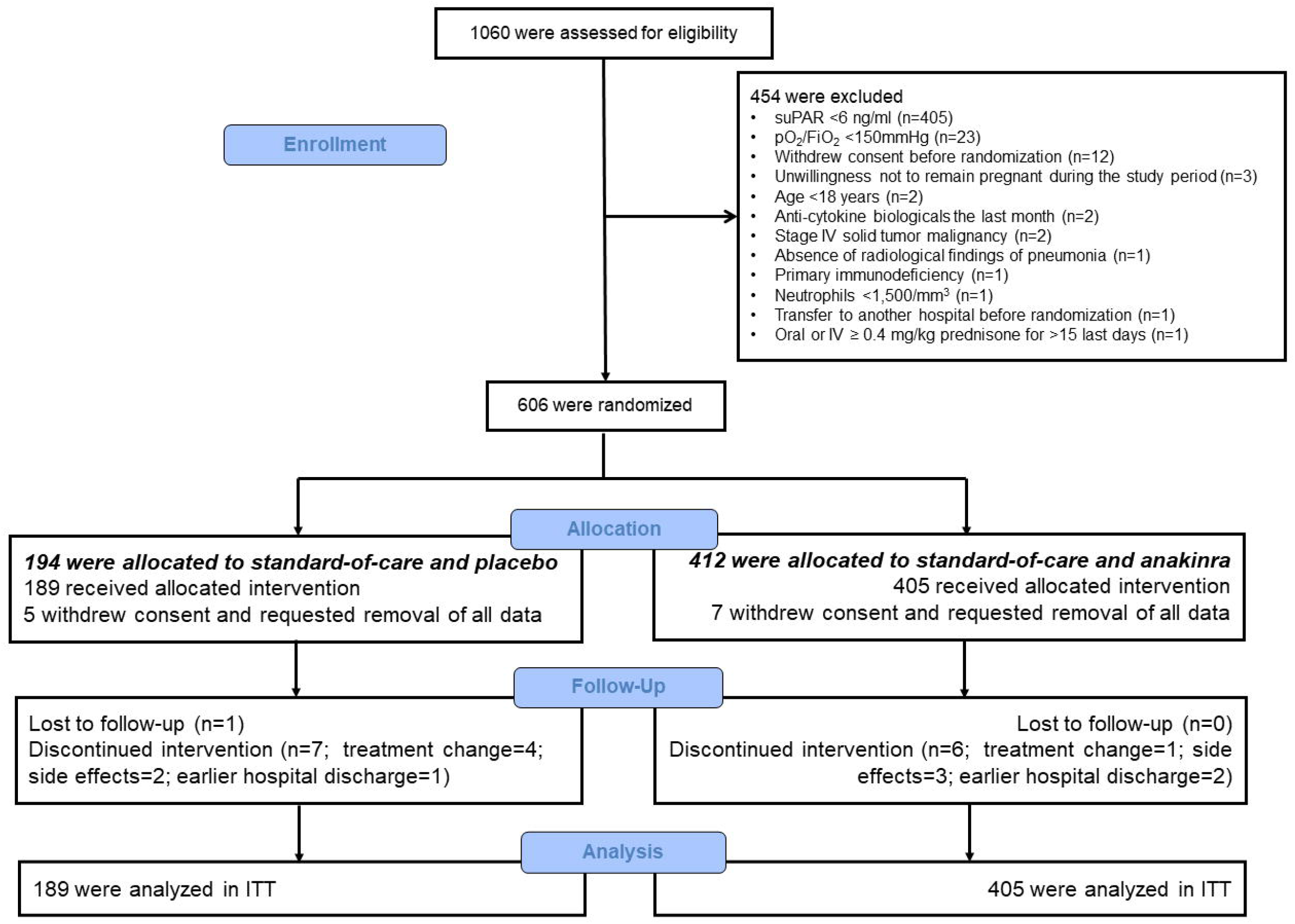
Study flow chart. <u>Abbreviations</u>: FiO_2_: fraction of inspired oxygen; ITT: intention-to-treat; IV: intravenous; pO_2_: partial oxygen pressure; suPAR: soluble urokinase plasminogen activator receptor

**Table 1.** Baseline Characteristics of Enrolled Patients.

|  | **SoC + Placebo**<br>**(N=189)** | **SoC + Anakinra**<br>**(N=405)** | **All patients**<br>**(N=594)** |
| --- | --- | --- | --- |
| Age, years, mean (SD) | 61.5 (11.3) | 62.0 (11.4) | 61.9 (12.1) |
| Male sex, n (%) | 108 (57.1) | 236 (58.3) | 344 (57.9) |
| Mean body mass index (SD) | 29.8 (5.6) | 29.4 (5.5) | 29.5 (5.5) |
| Charlson's comorbidity index, mean (SD) | 2.2 (1.5) | 2.3 (1.6) | 2.2 (1.6) |
| SOFA score, mean (SD) | 2.5 (1.2) | 2.4 (1.1) | 2.4 (1.1) |
| WHO classification for COVID-19, n (%) |  |  |  |
| Moderate pneumonia | 27 (14.3) | 82 (20.2) | 109 (18.4) |
| Severe pneumonia | 162 (85.7) | 323 (79.8) | 485 (81.6) |
| Days to start of study drug, median (Q1-Q3) |  |  |  |
| From symptom onset | 9 (7-11) | 9 (7-12) | 9 (7-11) |
| From hospital admission | 2 (2-3) | 2 (2-3) | 2 (2-3) |
| Laboratory values, median (Q1-Q3) |  |  |  |
| White blood cell count, cells per mm<sup>3</sup> | 5910 (4280-8300) | 5980 (4320-8180) | 5950 (4310-8200) |
| Lymphocyte count, cells per mm<sup>3</sup> | 730 (560-1090) | 815 (570-1110) | 800 (565-1100) |
| C-reactive protein, mg/l | 51.4 (25.2-98.5) | 50.5 (25.2-100.2) | 50.6 (25.3-99.7) |
| Interleukin-6, pg/ml | 20.1 (7.4-45.0) | 15.5 (6.7-39.3) | 16.8 (7.0-39.8) |
| Ferritin, ng/ml | 628.6<br>(293.5-1062.3) | 558.9<br>(294.1-1047.0) | 585.2<br>(294.5-1047.0) |
| Serum soluble uPAR, ng/ml | 7.5 (6.9-9.3) | 7.6 (7.0-9.1) | 7.6 (6.9-9.1) |
| PaO<sub>2</sub>: FiO<sub>2</sub> | 215 (161-293) | 235 (178-304) | 230 (172-300) |
| Comorbidities, no. (%) |  |  |  |
| Type 2 diabetes mellitus | 28 (14.8) | 66 (16.3) | 94 (15.8) |
| Chronic heart failure | 5 (2.6) | 13 (3.2) | 18 (3.0) |
| Chronic renal disease | 1 (0.5) | 9 (2.2) | 10 (1.7) |
| Chronic obstructive pulmonary disease | 9 (4.8) | 15 (3.7) | 24 (4.0) |
| Coronary heart disease | 13 (6.9) | 28 (6.9) | 41 (6.9) |
| Atrial fibrillation | 8 (4.2) | 20 (4.9) | 28 (4.7) |
| Depression | 9 (4.8) | 25 (6.2) | 34 (5.7) |
| Administered doses of study drug, mean (SD) | 8.7 (2.0) | 8.4 (2.1) | 8.6 (1.8) |
| Co-administered medications, n (%) |  |  |  |
| Remdesivir | 133 (70.4) | 294 (72.6) | 427 (71.9) |
| Dexamethasone at enrolment | 160 (84.7) | 326 (80.5) | 486 (81.8) |
| Dexamethasone over follow-up due to progression from moderate to severe disease | 8 (4.2) | 16 (4.4) | 26 (4.4) |
| Low molecular weight heparin | 175 (92.6) | 385 (95.1) | 560 (94.3) |
| β-lactamases | 10 (5.3) | 23 (5.7) | 33 (5.6) |
| Piperacillin/tazobactam | 36 (119.0) | 64 (15.8) | 100 (16.8) |
| Ceftriaxone | 85 (45.0) | 155 (38.3) | 240 (40.4) |
| Ceftaroline | 32 (16.9) | 75 (18.5) | 107 (18.0) |
| Respiratory fluoroquinolone | 24 (12.7) | 53 (13.1) | 77 (13.0) |
| Azithromycin | 35 (18.5) | 76 (18.8) | 111 (18.7) |
| Any glycopeptide | 19 (10.1) | 24 (5.9) | 43 (7.2) |
| Linezolid | 22 (11.6) | 45 (11.1) | 67 (11.3) |

Abbreviations: FiO<sub>2</sub>: fraction of inspired oxygen; PaO<sub>2</sub>: partial oxygen pressure; SD standard deviation; SOFA sequential organ failure assessment.; Q quartile; WHO World Health Organization

### Primary and secondary outcomes

The unadjusted OR of the WHO-CPS by day 28 was 0.36 (95%CI 0.26-0.49; P<0.001) (Figure 2A and Table 2) corresponding to 2.78 times better improvement of the clinical status. The testing of the assumptions of the ordinal regression analysis i.e. the Goodness-of-fit test and the parallel lines test were not statistically significant denoting an even distribution of the treatment effect size for all 11-points of the WHO-CPS. At the univariate analysis treatment with anakinra and dexamethasone and disease severity were significantly associated with the final outcome. However, in the multivariate analysis, treatment with anakinra was the only variable that was significantly associated with final outcome (OR 0.36; 95%CI 0.26-0.50; P<0.001) (Figure 2B).

**Figure 2.**
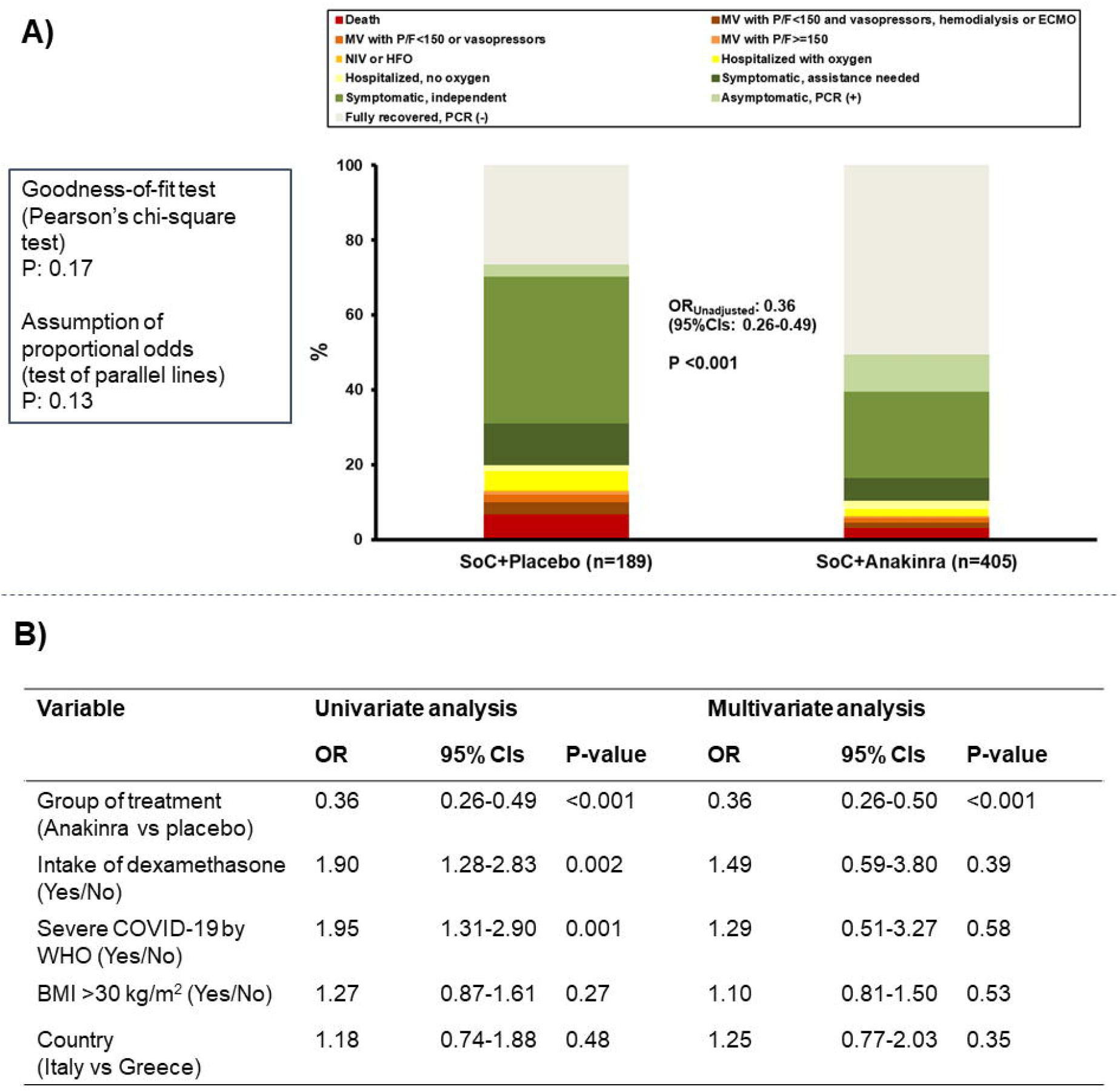
Study Primary Outcome. **A)** Distribution of the World Health Organization (WHO) Clinical Progression Scale (CPS) at day 28 of patients allocated to treatment with standard-of-care (SoC) and placebo and to treatment with SoC and anakinra. The odds ratio (OR) of the unadjusted ordinal regression analysis and the 95% confidence intervals (CIs) are shown. The two tests of the assumptions of the ordinal regression analysis are also provided. **B)** Univariate and multivariate ordinal regression analysis of the WHO-CPS at day 28. Co-variates entered in the multivariate model were those used for stratified randomization according to the received advice by the COVID-ETF of the EMA. <u>Abbreviations</u> CI: confidence interval; ECMO: extracorporeal membrane oxygenation; HFO: high flow oxygen; MV: mechanical ventilation; NIV: non-invasive ventilation; OR: odds ratio; PCR: polymerase chain reaction; P/F: respiratory failure; SoC; standard-of-care

**Table 2.** Primary and Secondary Study Outcomes.

|  | **SoC + Placebo**<br>**(N=189)** | **SoC + Anakinra**<br>**(N=405)** | **Odds Ratio**<br>**(95% CI)** | **P Value** |
| --- | --- | --- | --- | --- |
| WHO-CPS by day 28 |  |  | 0.36 (0.26-0.49) | <0.001 |
| Fully recovered PCR(-), n (%) | 50 (26.5) | 204 (50.4) |  |  |
| Asymptomatic PCR (+), n (%) | 6 (3.2) | 40 (9.9) |  |  |
| Symptomatic independent, n (%) | 74 (39.2) | 93 (23.0) |  |  |
| Symptomatic assistance needed, n (%) | 21 (11.1) | 25 (6.2) |  |  |
| Hospitalized no need for oxygen, n (%) | 3 (1.6) | 9 (2.2) |  |  |
| Hospitalized with nasal/mask oxygen, n (%) | 10 (5.3) | 8 (2.0) |  |  |
| Need for HFO or NIV, n (%) | 1 (0.5) | 1 (0.2) |  |  |
| Mechanical ventilation with P/F >150, n (%) | 1 (0.5) | 1 (0.2) |  |  |
| Mechanical ventilation with P/F <150 or vasopressors, n (%) | 4 (2.1) | 5 (1.2) |  |  |
| Mechanical ventilation with P/F <150 and vasopressors or<br>hemodialysis or ECMO, n (%) | 6 (3.2) | 6 (1.5) |  |  |
| Dead, n (%) | 13 (6.9) | 13 (3.2) |  |  |
| Absolute decrease of WHO-CPS at day 28 from baseline day 1, median (IQR) | 3 (2.5) | 4 (2.0) | 0.40 (0.29-0.55) | <0.001 |
| Absolute decrease of WHO-CPS at day 14 from baseline day 1, median (IQR) | 2 (3.0) | 3 (2.0) | 0.63 (0.46-0.85) | 0.003 |
| Absolute decrease of SOFA score at day 7 from baseline day 1, median (IQR) | 0 (1) | 1 (2) | 0.63 (0.46-0.86) | 0.004 |
| Median (IQR) time to hospital discharge, days | 12 (8.5) | 11 (7.8) | 1.22 (1.02-1.47)** | 0.033 |
| Median (IQ) time of ICU stay, days* | 14 (22) | 10 (21) | 2.33 (1.11-4.92)** | 0.026 |

\*only for patients admitted in the ICU
\*\*Hazard ratio
Abbreviations CI: confidence interval; ECMO: extra corporeal membrane oxygenation; HFO: high-flow oxygen; ICU: intensive care unit; IQR: interquartile range; MV: mechanical ventilation; NIV: non-invasive ventilation; PCR: polymerase chain reaction; P/F: respiratory ratio; SOFA: sequential organ failure assessment; WHO-CPS; World Health Organization Clinical Progression Scale

The three confirmatory analyses fully supported the clinical benefit of anakinra treatment. More precisely, the unadjusted OR of the ordinal regression analysis of the WHO-CPS by day 14 was 0.57 (95% CI 0.42-0.77; P<0.001) (Figure S1); after multivariate adjustment this was 0.58 (0.42-0.79; P: 0.001) (Table S1) showing also that anakinra treatment was the only variable affecting the outcome of patients by day 14. Regarding the second confirmatory analysis the multivariate logistic regression model for the first spectrum of the WHO-CPS showed that anakinra treatment and baseline severity were associated with persistence of the disease by day 28; anakinra was protective from disease persistence (OR: 0.36; 95% CI 0.25-0.53; P<0.001) (Table S2). The multivariate logistic regression model for the second spectrum of the WHO-CPS showed that anakinra treatment was the only variable that was associated with severe disease or death; anakinra was protective (OR: 0.46; 95% CI 0.26-0.83; P: 0.010) (Table S2). 28-day mortality was also lower among patients allocated to SoC and anakinra treatment (6.9% versus 3.2% respectively) (Figure S2). The third confirmatory analysis validated the results of the phase 2 SAVE trial. In this analysis, anakinra treatment prevented the progression to respiratory failure by day 14 (Figure S3 and Table S3) (31.7% in the SoC and placebo arm versus 20.7% in the SoC and anakinra arm).

The rate of protocol deviations from the SoC treatment was significantly greater among patients allocated to the placebo arm than patients allocated to the anakinra arm (14.3% versus 3.2% respectively; P<0.001). These protocol deviations in the SoC and placebo arm were mainly related to increasing the dose and/or duration of dexamethasone administration (Table S4). All five sensitivity analyses confirmed further the analysis of the primary endpoint (Table S5).

Analysis of the five clinical secondary endpoints showed a significant benefit from anakinra treatment on all of these endpoints. More precisely, the decreases of the WHO-CPS score from baseline by days 28 and 14 and of the SOFA score from baseline by day 7 were significantly greater in the SoC and anakinra arm (Table 2 and Tables S6 to S8). Moreover, in the anakinra group, the average time until hospital discharge was one 1 day shorter and the time until ICU discharge was 4 days shorter (Table 2 and Figures S4 and S5).

Over-time follow-up of laboratory values showed that among patients treated with anakinra: a) the absolute lymphocyte count was increased by day 7; b) circulating IL-6 was decreased by days 4 and 7; and c) plasma C-reactive protein (CRP) was decreased by day 7 (Figure S6).

### Adverse events

Overall, the incidence of serious TEAEs through day 28 was lower in patients in the anakinra and SoC group (16.5%) compared to the placebo and SoC group (21.2%). The non-serious TEAEs were similar in both treatment groups (Table 3 and Tables S9 and S10).

**Table 3.** Most common (>2%) Serious and non-Serious Treatment-Emergent Adverse Events (TEAE)

|  | **SoC+ Placebo**<br>**(n=189)** | **SoC+ Anakinra**<br>**(n=405)** | **P-value** |
| --- | --- | --- | --- |
| At least one serious TEAE, n (%) | 41 (21.2) | 68 (16.5) | 0.17 |
| Type of serious TEAE, n (%) |  |  |  |
| Infections and infestations, total | 25 (13.0) | 31 (7.5) | 0.035 |
| Ventilator-associated pneumonia | 14 (7.4) | 14 (3.5) | 0.039 |
| Bloodstream infection | 6 (3.2) | 12 (3.0) | 1.00 |
| Probable nosocomial infections | 4 (2.1) | 10 (2.5) | 1.00 |
| Pulmonary embolism | 4 (2.1) | 7 (1.7) | 0.75 |
| At least one non-serious TEAE, n (%) | 170 (90.4) | 352 (87.8) | 0.40 |
| Type of adverse event — no. (%) |  |  |  |
| Neutropenia | 1 (0.5) | 12 (3.0) | 0.07 |
| Anemia | 37 (19.6) | 58 (14.3) | <0.001 |
| Thrombocytopenia | 4 (2.1) | 9 (2.2) | >0.99 |
| Rash at the injection site | 3 (1.5) | 15 (3.7) | 0.20 |
| Constipation | 16 (8.5) | 39 (9.6) | 0.76 |
| Diarrhea | 8 (4.2) | 14 (3.5) | 0.82 |
| Increase of liver function tests | 63 (33.3) | 145 (35.8) | 0.58 |
| Bradycardia | 19 (10.1) | 36 (8.9) | 0.76 |
| Headache | 8 (4.2) | 16 (4.0) | >0.99 |
| Anxiety | 11 (5.8) | 33 (8.2) | 0.40 |
| Creatinine increase | 9 (4.8) | 17 (4.2) | 0.83 |
| Hyperglycemia | 76 (40.2) | 148 (36.5) | 0.41 |
| Hyponatremia | 23 (12.2) | 32 (7.9) | 0.13 |
| Hypernatremia | 17 (9.0) | 46 (11.4) | 0.40 |
| Hypokalemia | 12 (6.3) | 11 (2.7) | 0.04 |
| Hyperkalemia | 13 (6.9) | 36 (8.9) | 0.43 |
| Hypocalcemia | 20 (10.6) | 32 (7.9) | 0.35 |

Abbreviations: SoC: standard-of-care

## DISCUSSION

The SAVE-MORE trial showed an impressive efficacy of 10 days subcutaneous administration of anakinra in patients with COVID-19 and plasma suPAR 6 ng/ml or more. The global improvement of the overall clinical status was 2.78 times and it was evenly distributed in all scales of the day 28 WHO-CPS. The benefit was already apparent from day 14 and this is of major clinical importance since the first 14 days is the period during which a patient is expected to worsen; anakinra benefit was expanded until day 28. The magnitude of the efficacy of anakinra was shown in all multivariate analyses where in the presence of anakinra treatment the effect of disease severity on the final outcome was lost. The proportion of patients fully recovered exceeded 50% and those who remained under severe disease were reduced by 54%; the significant relative decrease of 28-day mortality was 55%. The large majority of the study population had severe COVID-19 and 81.8% were receiving SoC treatment containing dexamethasone. The remarkable improvement of patients under anakinra is also indirectly evidenced by the lack of changes in the SoC regimen. On the contrary, in 17% of patients receiving placebo, treating physicians changed the dexamethasone regimen and they administered either higher doses or even anti-cytokine biologicals.

The results fully validate the findings of the previous SAVE open-label phase II trial. In SAVE, the incidence of respiratory failure after 14 days with anakinra treatment was 22.3%^5^; in the SAVE-MORE trial it was 20.7%. For those who were eventually admitted to the ICU, time until discharge was significantly shorter in the anakinra and SoC treated group than in the placebo and SoC group; this was also observed in the previous SAVE trial^5^.

Early since the beginning of the COVID-19 pandemic, immunomodulators were suggested as one main strategy to attenuate the exaggerated immune response of the host. The most common administered drugs are anakinra and tocilizumab targeting the IL-1 and the IL-6 pathways respectively. However, the results of RCTs were heterogeneous and provided varying clinical benefit. There are nine published studies on the clinical efficacy of anakinra^5,7-14^; four have retrospective design, another four have prospective design using parallel comparators and only one is an RCT. Although most of these studies report mortality benefit, it is difficult to compare the findings to the results of the SAVE-MORE trial. The studies differ with regard to selection of patients, severity of illness and stage of the disease. Also duration of treatment, dose and route of administration was variable. So far, WHO-CPS was not studied as primary endpoint. Indeed, four of the studies were done in patients with critical illness with plasma levels of CRP and ferritin strongly exceeding the levels of the SAVE-MORE study population^7-10^.

What makes the difference between SAVE-MORE and the rest of RCTs and can explain the overwhelming efficacy of treatment? The reason is very likely patient stratification using suPAR as a biomarker of inflammation and diseases severity to select the patients most likely to benefit from anakinra treatment. Based on experimental studies and clinical evidence, early increase of suPAR is pointing towards excess release of DAMPs^3^. Predominant DAMPs are IL-1α that is released from the lung epithelium and calprotectin, which is subsequently exerting systemic effects through the production of IL-1β, designating suPAR as an important biomarker for excessive IL-1 bioactivity. Anakinra blocks both IL-1α and IL-1β by blocking their common receptor. The attenuation of the inflammatory responses by anakinra was shown by the decrease of IL-6 and of CRP circulating concentrations and by the increase of the absolute lymphocyte counts.

The clinical benefit of tocilizumab has been studied in six RCTs. In four of these RCTs, the patient populations were much similar to the population of the SAVE-MORE trial^15-18^. Clinical benefit from tocilizumab treatment was shown in only one of these four trials. On the opposite, most of clinical benefit from tocilizumab treatment was found in the other two trials, namely RECOVERY^19^ and REMAP-CAP^20^, with participants suffering from critical illness. Mortality was decreased from 35% with usual care to 31% in the RECOVERY trial^19^ whilst the median number of organ support-free days were increased from 0 days with usual care to 10 days with tocilizumab treatment in the REMAP-CAP trial^20^. The benefit of the more severe patients by tocilizumab may be explained by the biology of the critical illness. We have previously shown that circulating monocytes in critical COVID-19 present with complex immune dysregulation characterized by decreased efficiency for antigen-presentation and inappropriate maintenance of the potential for excess cytokine production: this dysregulation was restored upon exposure to tocilizumab^21^.

In conclusion, the SAVE-MORE trial showed that early start of treatment with anakinra and SoC guided by the biomarker suPAR in patients hospitalized with moderate and severe COVID-19 is leading to 2.78 times better improvement of the overall clinical status as expressed by the WHO-CPS. The frequency for full recovery is increased and the incidence of respiratory failure or death is decreased. This leads to shorter hospital stay. This finding is of outmost clinical importance and carries a major public health dimension, given the ICU overload during the COVID-19 pandemic, especially in countries with limited ICU capacity.

## Data Availability

Any request should be addressed to the Sponsor-Helleic Institute for the Study of Sepsis

## Acknowledgment

The authors wish to express their gratitude to the members of the Data Monitoring and Safety Committee Professors Michael Niederman, Jos WM van der Meer and Konrad Reinhart for their valuable contribution and for their critical review of the submitted manuscript.

## REFERENCES

1. Rovina N, Akinosoglou K, Eugen-Olsen J, Hayek S, Reiser J, Giamarellos-Bourboulis EJ. Soluble urokinase plasminogen activator receptor (suPAR) as an early predictor of severe respiratory failure in patients with COVID-19 pneumonia. Crit Care 2020; 24: 187.

2. Azam TU, Shadid HR, Blakely P, et al. Soluble urokinase receptor (suPAR) in COVID-19-related AKI. J Am Soc Nephrol 2020; 31: 2725–35.

3. Renieris G, Karakike E, Gkavogianni T, et al. IL-1 mediates tissue specific inflammation and severe respiratory failure In Covid-19: clinical and experimental evidence. medRxiv doi.org/10.1101/2021.04.09.2125519

4. Rodrigues TS, de Sá KSG, Ishimoto AY, et al. Inflammasomes are activated in response to SARS-CoV-2 infection and are associated with COVID-19 severity in patients. J Exp Med 2021; 218: e20201707.

5. Kyriazopoulou E, Panagopoulos P, Metallidis S, et al. An open label trial of anakinra to prevent respiratory failure in COVID-19. eLife 2021; 10: e66125.

6. https://www.who.int/publications/i/item/clinical-management-of-covid-19

7. Bozzi G, Mangioni D, Minoia F, et al. Anakinra combined with methylprednisolone in patients with severe COVID-19 pneumonia and hyperinflammation: An observational cohort study. J Allergy Clin Immunol 2021; 147: 561–6.

8. Cavalli G, Larcher A, Tomelleri A, et al. Interleukin-1 and interleukin-6 inhibition compared with standard management in patients with COVID-19 and hyperinflammation: a cohort study. Lancet Rheumatol 2021; e253–61.

9. Kooistra EJ, Waalders NJB, Grondman I, et al. Anakinra treatment in critically ill COVID-19 patients: a prospective cohort study. Crit Care 2020; 24: 688.

10. Cauchois R, Koubi M, Delarbre D, et al. Early IL-1 receptor blockade in severe inflammatory respiratory failure complicating COVID-19. Proc Natl Acad Sci U S A. 2020; 117: 18951–3.

11. Pontali E, Volpi S, Signori A, et al. Efficacy of early anti-inflammatory treatment with high doses of intravenous anakinra with or without glucocorticoids in patients with severe COVID-19 pneumonia. J Allergy Clin Immunol. 2021; 147: 1217–25.

12. Huet T, Beaussier H, Voisin O, et al. Anakinra for severe forms of COVID-19: a cohort study. Lancet Rheumatol 2020; 2: e393–e400.

13. CORIMUNO-19 Collaborative group. Effect of anakinra versus usual care in adults in hospital with COVID-19 and mild-to-moderate pneumonia (CORIMUNO-ANA-1): a randomised controlled trial. Lancet Respir Med 2021; 9: 295–304.

14. Balkhair A, Al-Zakwani I, Al Busaidi M, et al. Anakinra in hospitalized patients with severe COVID-19 pneumonia requiring oxygen therapy: Results of a prospective, open-label, interventional study. Int J Infect Dis 2021; 103: 288–96.

15. Rosas IO, Bräu N, Waters M, et al. Tocilizumab in hospitalized patients with severe Covid-19 pneumonia. N Engl J Med 2021; 384: 1503–16

16. Salama C, Han J, Yau L, Reiss WG, et al. Tocilizumab in patients hospitalized with Covid-19 pneumonia. N Engl J Med 2021; 384: 20–30.

17. Stone JH, Frigault MJ, Serling-Boyd NJ, et al. Efficacy of tocilizumab in patients hospitalized with Covid-19. N Engl J Med 2020; 383: 2333–44.

18. Salvarani C, Dolci G, Massari M, et al. Effect of Tocilizumab vs standard care on clinical worsening in patients hospitalized with COVID-19 pneumonia: a randomized clinical trial. JAMA Intern Med 2021; 181: 24–31.

19. RECOVERY collaborative group. Tocilizumab in patients admitted to hospital with COVID-19 (RECOVERY): a randomised, controlled, open-label, platform trial. Lancet 2021; 397: 1637–45.

20. Gordon AC, Mouncey PR, Al-Beidh F, et al. Interleukin-6 receptor antagonists in critically ill patients with Covid-19. N Engl J Med 2021; 384: 1491–1502.

21. Giamarellos-Bourboulis EJ, Netea MG, Rovina N, et al. Complex immune dysregulation in COVID-19 patients with severe respiratory failure. Cell Host Microbe 2020; 27: 992–1000

