## Supplement for "Early Anakinra Treatment for COVID-19 Guided by Urokinase Plasminogen Receptor"

**SUPPLEMENTARY MATERIAL FOR THE SAVE-MORE TRIAL**

**Table of contents**

| **Content** | **Page** |
| --- | --- |
| Supplementary Figures | 3 |
| Supplementary Figure 1. First supporting analysis of the study primary outcome. | 3 |
| Supplementary Figure 2. Survival analysis of enrolled patients | 4 |
| Supplementary Figure 3. Time to progression into severe respiratory failure | 5 |
| Supplementary Figure 4. Time to hospital discharge. | 6 |
| Supplementary Figure 5. Time to discharge from the Intensive Care Unit | 7 |
| Supplementary Figure 6. Levels of lymphocytes, interleukin (IL)-6 and C-reactive protein (CRP) over days of follow-up | 8 |
| Supplementary tables | 9 |
| Supplementary Table 1. First confirmatory analysis of the primary endpoint | 9 |
| Supplementary Table 2. Second confirmatory analyses of the primary endpoint | 10 |
| Supplementary Table 3. Third confirmatory analysis of the primary endpoint | 12 |
| Supplementary Table 4. Recorded deviations from the per-protocol standard-of-care treatment | 13 |
| Supplementary Table 5. The five sensitivity analyses for the primary study endpoint | 15 |
| Supplementary Table 6. Changes of the World Health Organization Clinical Progression Scale (WHO-CPS) at day 28 from baseline | 19 |
| Supplementary Table 7. Changes of the World Health Organization Clinical Progression Scale (WHO-CPS) by day 14 from baseline | 20 |
| Supplementary Table 8. Changes of the Sequential Organ Failure Assessment score at day 7 from baseline | 21 |
| Supplementary Table 9. Complete list of serious treatment-emergent adverse events (TEAE) Classified by System | 22 |
| Supplementary Table 10. Complete list of non-serious treatment-emergent adverse events (TEAE) Classified by System | 26 |

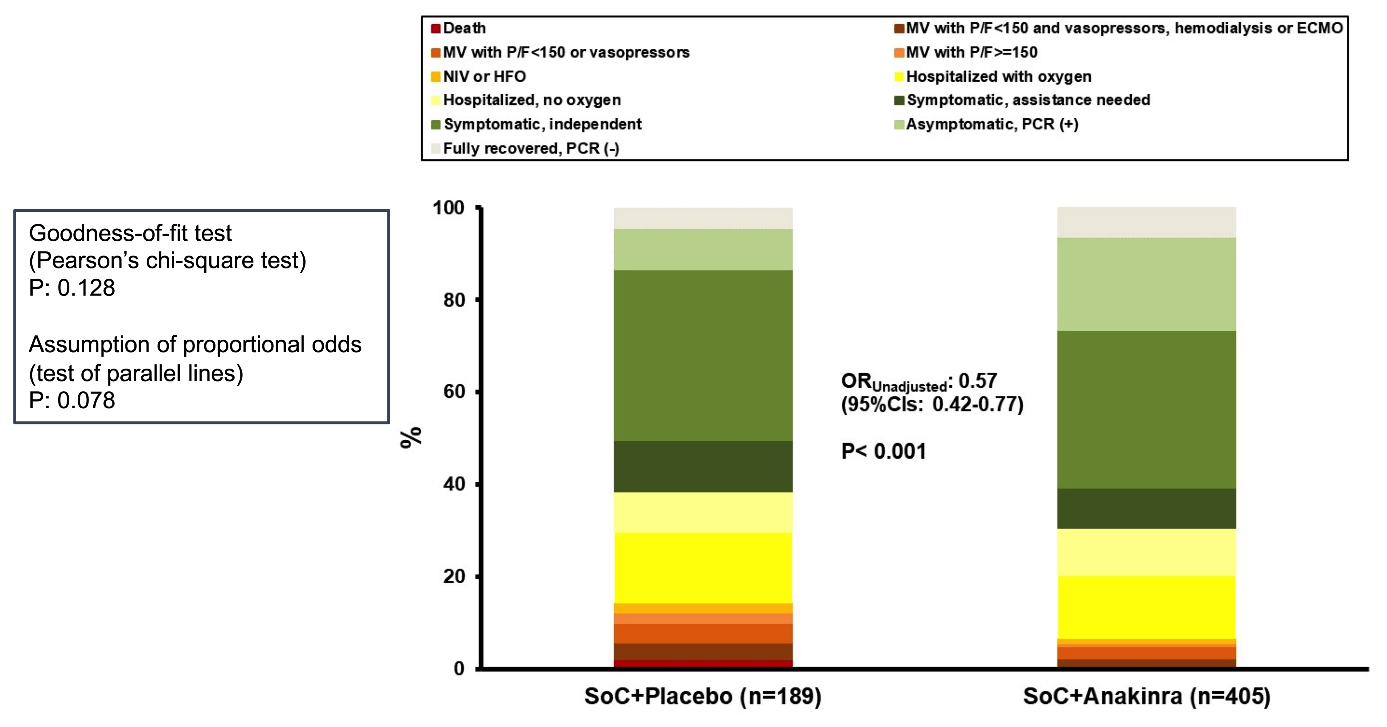

**Figure S1. First supporting analysis of the study primary endpoint**

Distribution of the World Health Organization (WHO) Clinical Progression Scale (CPS) at day 14 of patients allocated to treatment with standard-of-care (SoC) and placebo and to treatment with SoC and anakinra. The odds ratio (OR) of the unadjusted ordinal regression analysis and the 95% confidence intervals (CIs) are shown. The two tests of the assumptions of the ordinal regression analysis are also provided.

Abbreviations CI: confidence interval; ECMO: extracorporeal membrane oxygenation; HFO: high flow oxygen; MV: mechanical ventilation; NIV: non-invasive ventilation; OR: odds ratio; PCR: polymerase chain reaction; P/F: respiratory failure; SoC; standard-of-care

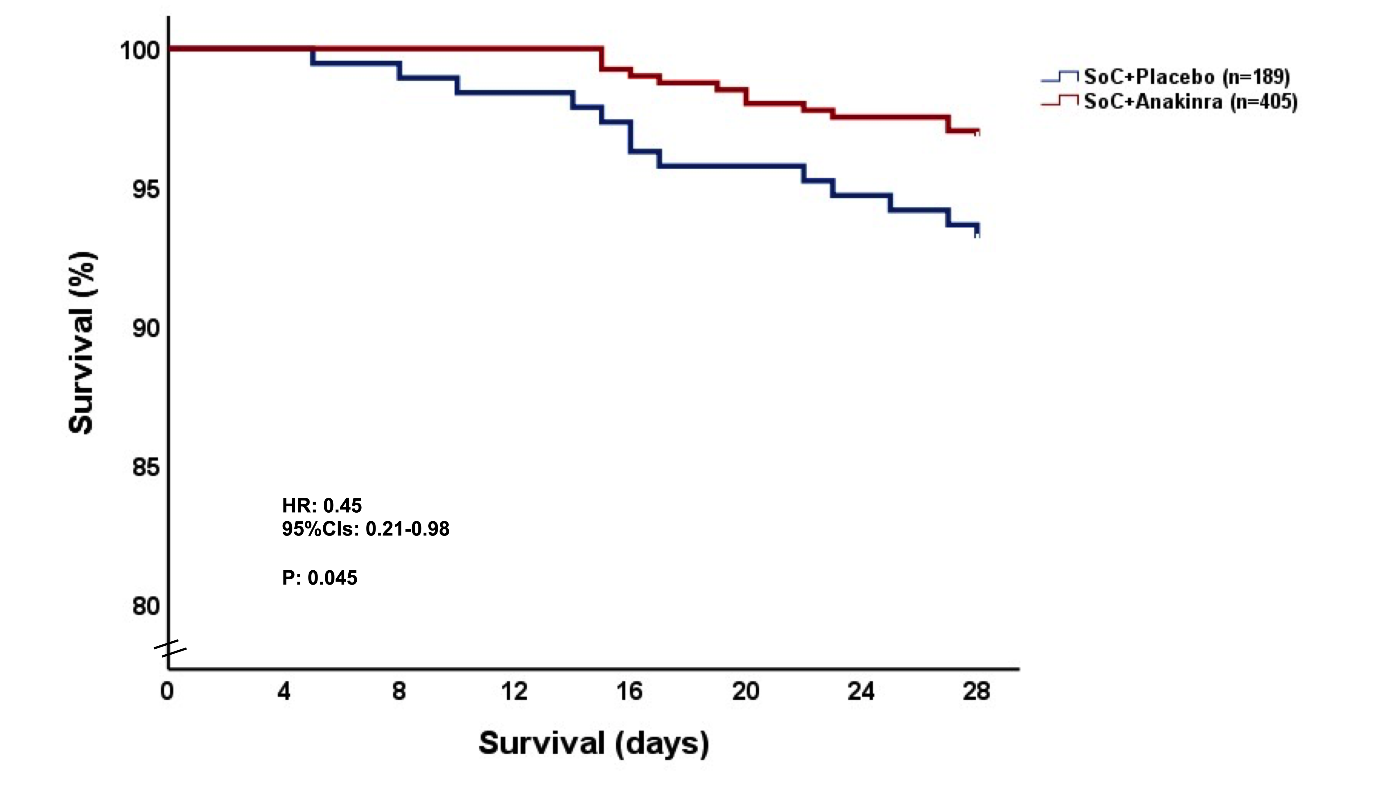

**Figure S2 Survival analysis of enrolled patients**

Abbreviations: CI: confidence intervals; HR: hazard ratio; SoC: standard-of-care

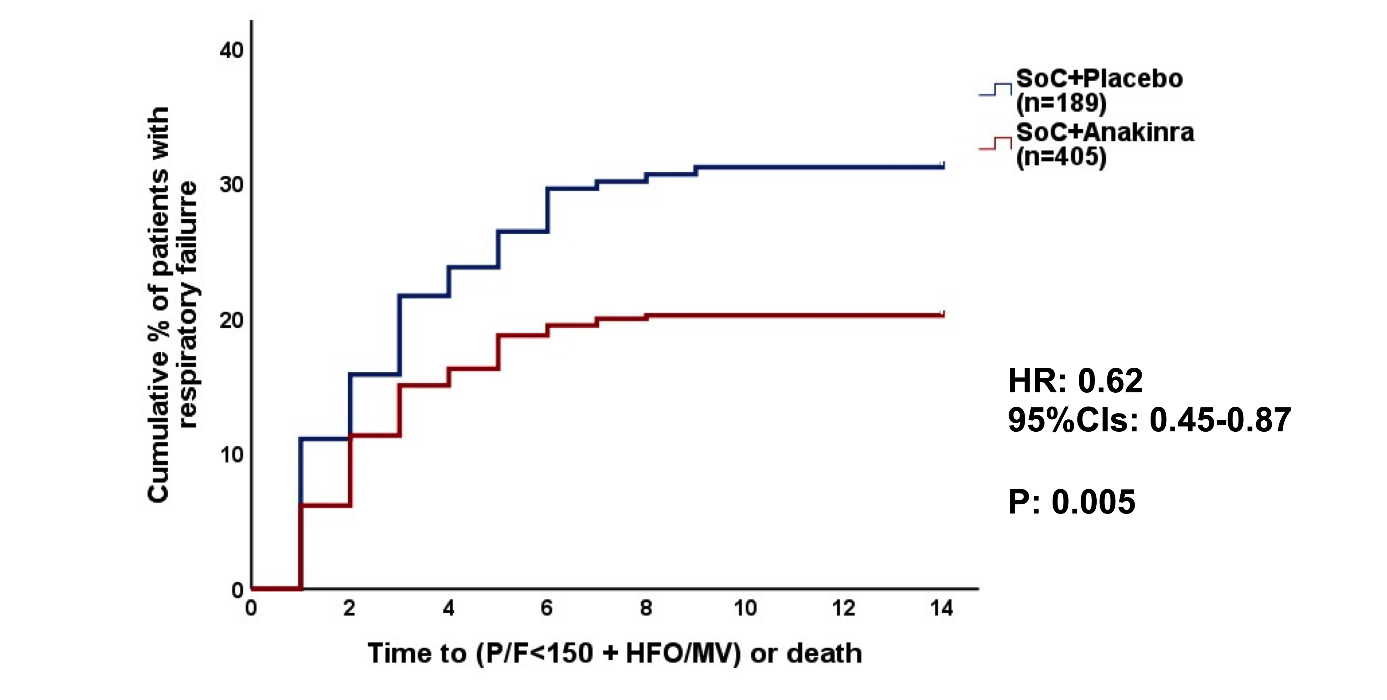

**Figure S3** **Time to progression into severe respiratory failure**

Respiratory failure is defined as respiratory ratio-PF<150 necessitating high flow oxygen/non-invasive ventilation/mechanical ventilation or death) by day 14. The hazard ratio (HR) and the 95% confidence intervals (CIs) are provided.

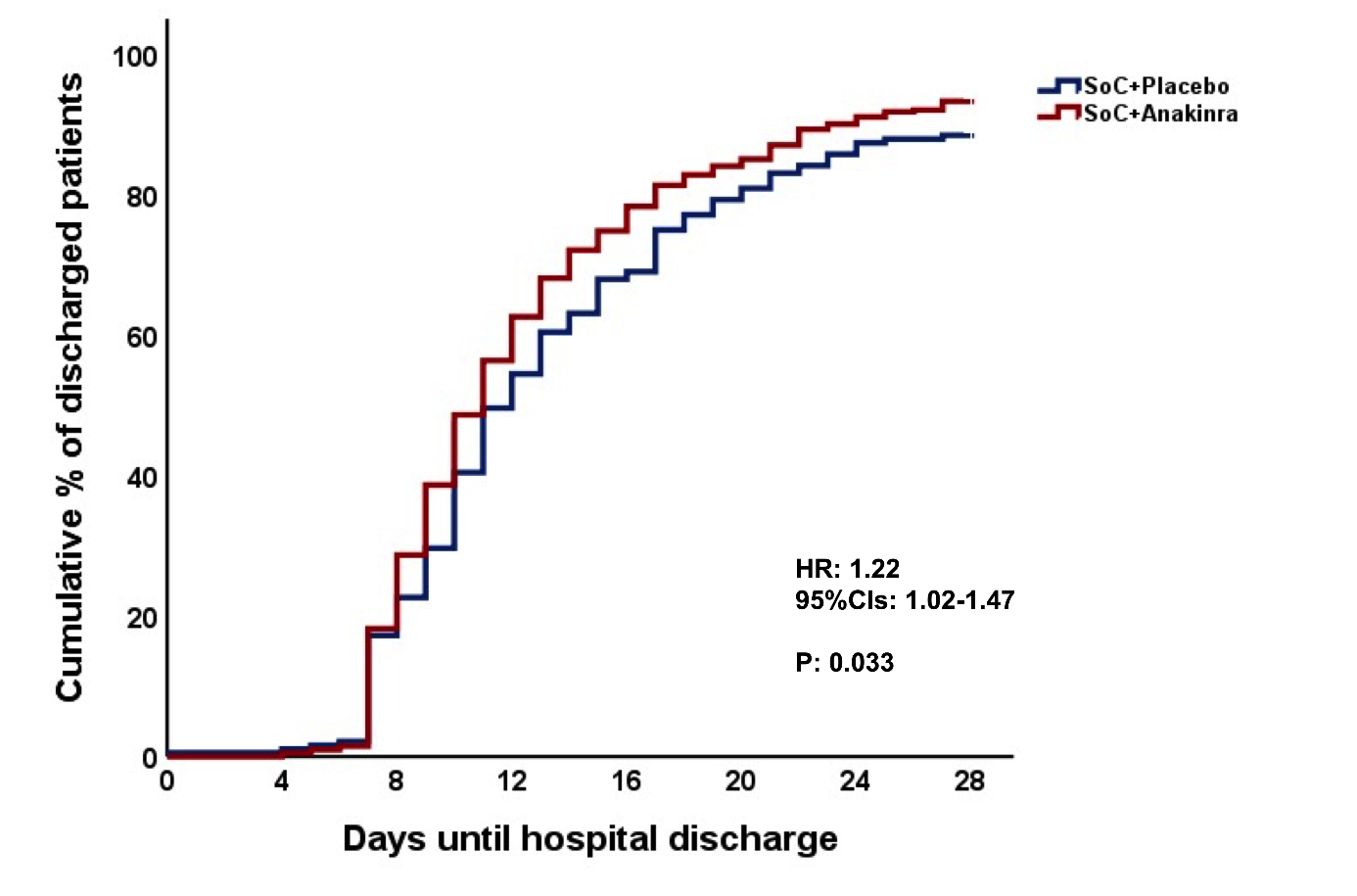

**Figure S4 Time to hospital discharge**

Abbreviations: CI: confidence intervals; HR: hazard ratio; SoC: standard-of-care

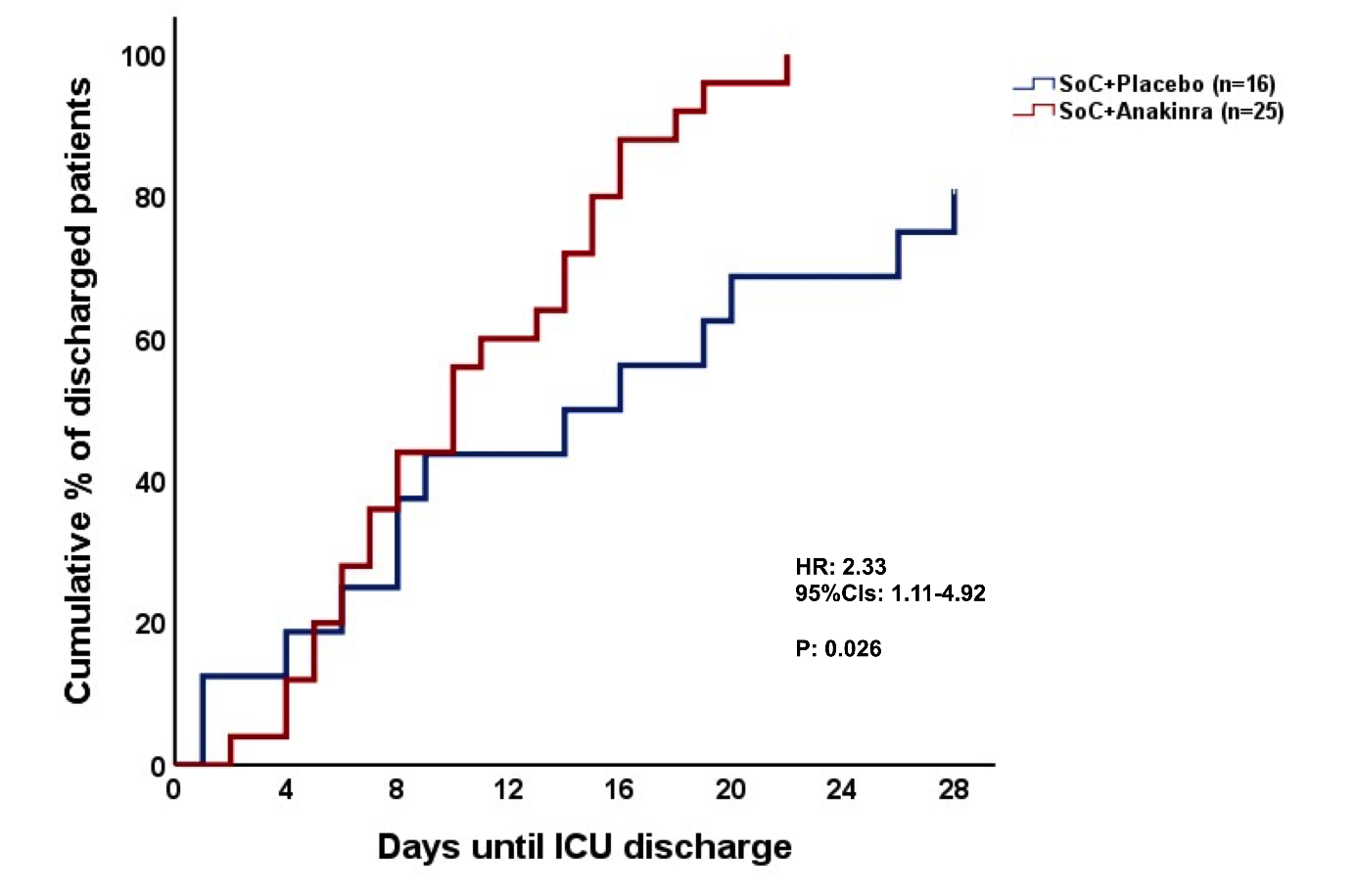

**Figure S5 Time to discharge from the intensive care unit**

Analysis involves only patients who were admitted in the intensive care unit

Abbreviations: CI: confidence intervals; HR: hazard ratio; SoC: standard-of-care

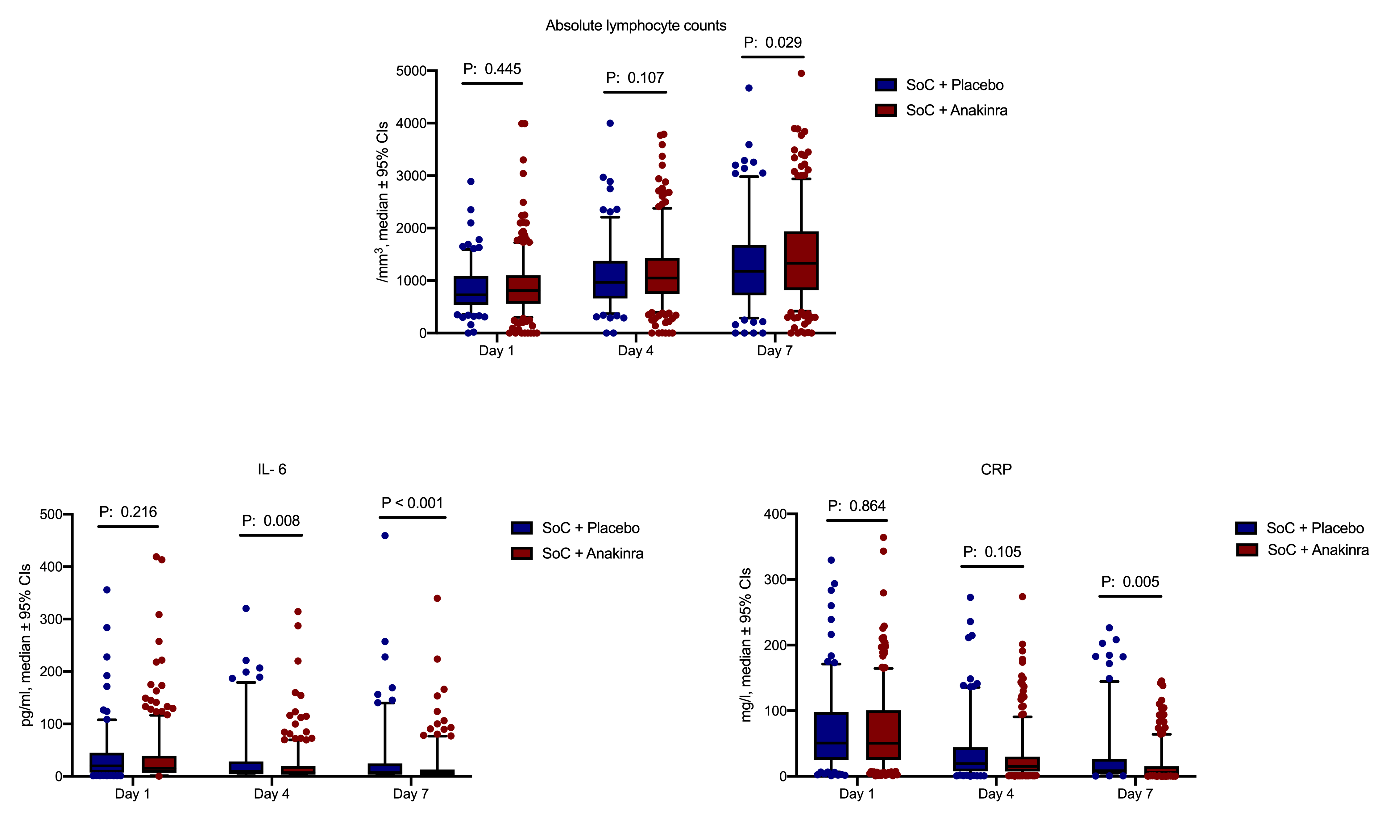

**Figure S6 Levels of lymphocytes, interleukin (IL)-6 and C-reactive protein (CRP) over days of follow-up**

Day 1 sampling was done before start of administration of the study drug. The P-values of comparisons for each day of follow-up are provided

Abbreviations: CI: confidence interval; SoC: standard-of-care

**Table S1** **First confirmatory analysis of the primary endpoint**

Univariate and multivariate ordinal regression analysis of the primary study outcome (World Health Organization Clinical Progression Scale) at day 14. Co-variates entered in the multivariate model were those used for stratified randomization according to the received advice by the COVID-ETF of the EMA.

| **Variable** | **Univariate analysis** | | | **Multivariate analysis** | | |
| --- | --- | --- | --- | --- | --- | --- |
|  | Odds ratio | 95% CIs | P-value | Odds ratio | 95% CIs | P-value |
| Group of treatment (Anakinra vs placebo) | 0.57 | 0.42-0.77 | <0.001 | 0.58 | 0.42-0.79 | <0.001 |
| Intake of dexamethasone (Yes/No) | 2.23 | 1.53-3.26 | <0.001 | 1.69 | 0.70-4.11 | 0.24 |
| Severe COVID-19 by WHO (Yes/No) | 2.23 | 1.53-3.26 | <0.001 | 1.36 | 0.56-3.27 | 0.49 |
| BMI >30 kg/m^2^ (Yes/No) | 1.15 | 0.86-1.56 | 0.34 | 1.05 | 0.78-1.42 | 0.72 |
| Country  (Italy vs Greece) | 1.14 | 0.72-1.79 | 0.57 | 1.26 | 0.79-2.00 | 0.32 |

Abbreviations BMI: body mass index; CI: confidence internal

**Table S2 Second confirmatory analyses of the primary endpoint**

Univariate and multivariate step-wise logistic regression analysis of the primary study outcome (World Health Organization Clinical Progression Scale-WHO-CPS) at day 28 towards the two spectra of this scale. The first spectrum involves patients with fully resolved disease (0 points of the WHO-CPS) or persistent disease (1 to 10 points of the WHO-CPS). The second spectrum involves patients with severe disease or dead (6 to 10 points of the WHO-CPS) or not that severe (0 to 5 points of the WHO-CPS) Co-variates entered in the multivariate model were those used for stratified randomization according to the received advice by the COVID-ETF of the EMA.

| **Variable** |  |  | **Univariate analysis** | | | **Multivariate analysis** | |
| --- | --- | --- | --- | --- | --- | --- | --- |
|  | **Analysis towards fully resolved or persistent disease** | | | | | | |
|  | **Fully resolved**  **(n= 254)** | **Persistence**  **(n= 340)** | **Odds ratio**  **(95% CIs)** | **P-value** | **Odds ratio**  **(95% CIs)** | | **P-value** |
| Anakinra treatment, n (%) | 204 (80.3) | 201 (59.1) | 0.35 (0.23-0.52) | <0.001 | 0.36 (0.25-0.53) | | <0.001 |
| Intake of dexamethasone, n (%) | 198 (78.0) | 288 (84.7) | 1.56 (1.03-2.38) | 0.036 | * | |  |
| Severe COVID-19 by WHO, n (%) | 196 (77.2) | 289 (85.0) | 1.68 (1.10-2.55) | 0.015 | 1.58 (1.02-2.42) | | 0.037 |
| BMI >30 kg/m^2^, n (%) | 87 (34.3) | 129 (37.9) | 1.17 (0.84-1.65) | 0.36 | * | |  |
| Patients in Italy, n (%) | 30 (11.8) | 37 (10.9) | 0.91 (0.55-1.52) | 0.72 | * | |  |
|  | **Analysis towards allocation into WHO-CPS ≥6 (Yes) or WHO-CPS ≤5** | | | | | | |
|  | **WHO-CPS ≤5 (n= 544)** | **WHO-CPS ≥6 (n= 50)** | **Odds ratio**  **(95% CIs)** | **P-value** | **Odds ratio**  **(95% CIs)** | | **P-value** |
| Anakinra treatment, n (%) | 379 (69.8) | 26 (50.1) | 0.45  (0.25-0.80) | 0.007 | 0.46  (0.26-0.83) | | 0.010 |
| Intake of dexamethasone, n (%) | 438 (80.1) | 50 (100) | * | <0.001 | * | |  |
| Severe COVID-19 by WHO, n (%) | 435 (80.0) | 50  (100) | * | <0.001 | ** | |  |
| BMI >30 kg/m^2^ n (%) | 200 (36.8) | 16 (32.0) | 0.81 (0.44-1.50) | 0.81 | ** | |  |
| Patients in Italy, n (%) | 59 (10.8) | 8 (16.0) | 1.57 (0.70-3.49) | 0.27 | ** | |  |

*cannot be computed because one value is zero

**variables not included in equation after 2 steps of forward analysis

Abbreviations BMI: body mass index; CI: confidence internal

**Table S3** **Third confirmatory analysis of the primary endpoint**

Univariate and multivariate step-wise Cox regression analysis of progression into respiratory failure the first 14 days. This analysis is done to validate the results of the phase 2 study SAVE. Respiratory failure as any respiratory ratio less than 150 requiring the use of high-flow oxygen/non-invasive ventilation/mechanical ventilation or death. Co-variates entered in the multivariate model were those used for stratified randomization according to the received advice by the COVID-ETF of the EMA.

| **Variable** | **Respiratory failure** | | **Univariate analysis** | | **Multivariate analysis** | |
| --- | --- | --- | --- | --- | --- | --- |
|  | **No**  **(n= 450)** | **Yes**  **(n= 144)** | **Hazard ratio**  **(95% CIs)** | **P-value** | **Hazard ratio**  **(95% CIs)** | **P-value** |
| Anakinra treatment, n (%) | 321 (71.3) | 84 (58.3) | 0.62 (0.45-0.87) | 0.005 | 0.67 (0.47-0.93) | 0.017 |
| Intake of dexamethasone, n (%) | 345 (76.7) | 141 (97.9) | 11.63 (3.70-36.49) | <0.001 | * |  |
| Severe COVID-19 by WHO n (%) | 343 (76.2) | 152 (98.6) | 18.27 (4.52-73.77) | <0.001 | 17.16 (4.25-69.33) | <0.001 |
| BMI >30 kg/m^2^, n (%) | 158 (35.1) | 58 (40.3) | 0.84 (0.60-1.17) | 0.31 | * |  |
| Patients in Italy, n (%) | 39 (8.7) | 28 (19.4) | 2.34 (1.55-3.54) | <0.001 | 2.21 (1.46-3.34) | <0.001 |

*variables not included in equation after 3 steps of forward analysis

Abbreviations BMI: body mass index; CI: confidence internal

**Table S4 Recorded deviations from the per-protocol standard-of-care treatment**

| **Deviation, n (%)** | **SoC + Placebo (n=189)** | **SoC + Anakinra (n= 405)** | **P-value** |
| --- | --- | --- | --- |
| Administration of dexamethasone for more than 10 days | 9 (4.8) | 0 (0) | <0.001 |
| Daily dosing of dexamethasone 10-20mg | 3 (1.6) | 2 (0.5) | 0.33 |
| Administration of dexamethasone 6-18 mg daily with MTP | 4 (2.1) | 0 (0) | 0.010 |
| Stop of study drug, administration of TCZ + IVIG+ ANA | 3 (1.6) | 0 (0) | 0.031 |
| Administration of secukinumab | 1 (0.5) | 0 (0) | 0.32 |
| Dexamethasone administration i moderate disease | 2 (1.1) | 3 (0.7) | 0.66 |
| Dexamethasone administration for 11 days and co-administration of TCZ and MTP | 0 (0) | 1 (0.2) | >0.99 |
| Administration of dexamethasone for less than 10 days | 1 (0.5) | 1 (0.2) | 0.54 |
| Premature stop of study drug due to leukopenia, n (%) | 1 (0.5) | 1 (0.2) | 0.54 |
| Premature stop of study drug due to increase of aminotransferases | 1 (0.5) | 2 (0.5) | >0.99 |
| Premature stop of study drug due to earlier hospital discharge | 1 (0.5) | 2 (0.5) | >0.99 |
| Premature stop of study drug by the attending physicians after ICU admission | 1 (0.5) | 1 (0.2) | 0.54 |

Abbreviations: ANA: anakinra; ICU: intensive care unit; IVIG: intravenous γ-globulin; MTP: methylprednisolone; SoC: standard-of-care; TCZ: tocilizumab

**Table S5 The five sensitivity analyses for the primary study endpoint**

The first four sensitivity analyses are univariate and multivariate ordinal regression analyses of the primary study outcome (World Health Organization Clinical Progression Scale) at day 28. Co-variates entered in the multivariate model were those used for stratified randomization according to the received advice by the COVID-ETF of the EMA.

|  | **Univariate analysis** | | | **Multivariate analysis** | | |
| --- | --- | --- | --- | --- | --- | --- |
|  | **Sensitivity analysis 1: Per-protocol (SoC + placebo= 162; SoC + anakinra= 292)** | | | | | |
|  | **Odds ratio** | **95% CIs** | **P-value** | **Odds ratio** | **95% CIs** | **P-value** |
| Group of treatment (Anakinra vs placebo) | 0.34 | 0.24-0.48 | <0.001 | 0.35 | 0.25-0.48 | <0.001 |
| Intake of dexamethasone (Yes/No) | 1.72 | 1.15-2.59 | 0.009 | 1.42 | 0.47-4.20 | 0.53 |
| Severe COVID-19 by WHO (Yes/No) | 1.76 | 1.17-2.67 | 0.007 | 1.22 | 0.41-3.68 | 0.71 |
| BMI >30 kg/m^2^ (Yes/No) | 1.16 | 0.84-1.59 | 0.36 | 1.111 | 0.80-1.53 | 0.43 |
| Country (Italy vs Greece) | 1.05 | 0.65-1.71 | 0.82 | 1.15 | 0.70-1.88 | 0.58 |
|  | **Sensitivity analysis 2: Population receiving ≥7 doses of the study drug**  **(SoC + placebo= 177; SoC + anakinra= 382)** | | | | | |
|  | **Odds ratio** | **95% CIs** | **P-value** | **Odds ratio** | **95% CIs** | **P-value** |
| Group of treatment (Anakinra vs placebo) | 0.37 | 0.28-0.52 | <0.001 | 0.38 | 0.27-0.53 | <0.001 |
| Intake of dexamethasone (Yes/No) | 1.90 | 1.27-2.86 | 0.002 | 1.14 | 0.42-3.11 | 0.79 |
| Severe COVID-19 by WHO (Yes/No) | 2.03 | 1.36-3.05 | 0.001 | 1.70 | 0.63-4.58 | 0.29 |
| BMI >30 kg/m^2^ (Yes/No) | 1.19 | 0.88-1.64 | 0.26 | 1.11 | 0.80-1.53 | 0.51 |
| Country (Italy vs Greece) | 1.21 | 0.74-1.99 | 0.44 | 1.26 | 0.75-2.10 | 0.38 |
|  | **Sensitivity analysis 3: Complete cases analysis**  **(SoC + placebo= 188; SoC + anakinra= 405)** | | | | | |
|  | **Odds ratio** | **95% CIs** | **P-value** | **Odds ratio** | **95% CIs** | **P-value** |
| Group of treatment (Anakinra vs placebo) | 0.35 | 0.26-0.49 | <0.0001 | 0.36 | 0.26-0.49 | <0.001 |
| Intake of dexamethasone (Yes/No) | 1.91 | 1.28-2.84 | 0.001 | 1.51 | 0.59-2.83 | 0.39 |
| Severe COVID-19 by WHO (Yes/No) | 1.96 | 1.32-2.92 | 0.001 | 1.29 | 0.51-3.27 | 0.59 |
| BMI >30 kg/m^2^ (Yes/No) | 1.19 | 0.89-1.63 | 0.24 | 1.12 | 0.82-1.52 | 0.49 |
| Country (Italy vs Greece) | 1.21 | 0.75-1.95 | 0.43 | 1.27 | 0.78-2.08 | 0.32 |
|  | **Sensitivity analysis 4: Responder analysis treating missing values as failures**  **(SoC + placebo= 189; SoC + anakinra= 405)** | | | | | |
|  | **Odds ratio** | **95% CIs** | **P-value** | **Odds ratio** | **95% CIs** | **P-value** |
| Group of treatment (Anakinra vs placebo) | 0.35 | 0.25-0.48 | <0.001 | 0.36 | 0.26-0.49 | <0.001 |
| Intake of dexamethasone (Yes/No) | 1.92 | 1.29-2.85 | 0.001 | 1.49 | 0.59-3.79 | 0.39 |
| Severe COVID-19 by WHO (Yes/No) | 1.97 | 1.32-2.93 | 0.001 | 1.30 | 0.52-3.28 | 0.57 |
| BMI >30 kg/m^2^ (Yes/No) | 1.21 | 0.89-1.65 | 0.21 | 1.14 | 0.83-1.55 | 0.42 |
| Country (Italy vs Greece) | 1.15 | 0.72-1.84 | 0.55 | 1.21 | 0.74-1.96 | 0.44 |
|  | **Sensitivity analysis 5: Comparison of the unadjusted and the adjusted model** | | | | | |
|  | **Unadjusted** | |  | **Adjusted** | |  |
|  | **Odds ratio** | **95% CI** |  | **Odds ratio** | **95% CI** | **P-value** |
| Group of treatment (Anakinra vs placebo) | 0.36 | 0.26-0.49 |  | 0.36 | 0.25-0.50 | 0.54 |

Abbreviations BMI: body mass index; CI: confidence internal

**Table S6 Changes of the World Health Organization Clinical Progression Scale (WHO-CPS) at day 28 from baseline**

Univariate and multivariate ordinal regression analyses are shown. Co-variates entered in the multivariate model were those used for stratified randomization according to the received advice by the COVID-ETF of the EMA.

| **Variable** | **Univariate analysis** | | | **Multivariate analysis** | | |
| --- | --- | --- | --- | --- | --- | --- |
|  | **Odds ratio** | **95% CIs** | **P-value** | **Odds ratio** | **95% CIs** | **P-value** |
| Group of treatment (Anakinra vs placebo) | 0.40 | 0.29-0.55 | <0.001 | 0.40 | 0.29-0.55 | <0.001 |
| Intake of dexamethasone (Yes/No) | 1.00 | 0.69-1.46 | 0.96 | 0.94 | 0.38-2.31 | 0.89 |
| Severe COVID-19 by WHO (Yes/No) | 1.07 | 0.73-1.55 | 0.74 | 1.07 | 0.43-2.60 | 0.89 |
| BMI >30 kg/m^2^ (Yes/No) | 1.08 | 0.79-1.46 | 0.62 | 1.02 | 0.75-1.39 | 0.88 |
| Country (Italy vs Greece) | 1.64 | 1.03-2.64 | 0.036 | 1.65 | 1.02-2.67 | 0.040 |

Abbreviations BMI: body mass index; CI: confidence internal

**Table S7 Changes of the World Health Organization Clinical Progression Scale (WHO-CPS) by day 14 from baseline**

Univariate and multivariate ordinal regression analyses are shown. Co-variates entered in the multivariate model were those used for stratified randomization according to the received advice by the COVID-ETF of the EMA.

| **Variable** | **Univariate analysis** | | | **Multivariate analysis** | | |
| --- | --- | --- | --- | --- | --- | --- |
|  | **Odds ratio** | **95% CIs** | **P-value** | **Odds ratio** | **95% CIs** | **P-value** |
| Group of treatment (Anakinra vs placebo) | 0.63 | 0.46-0.85 | 0.003 | 0.63 | 0.46-0.86 | 0.003 |
| Intake of dexamethasone (Yes/No) | 1.28 | 0.88-1.85 | 0.20 | 1.19 | 0.49-2.88 | 0.69 |
| Severe COVID-19 by WHO (Yes/No) | 1.30 | 0.90-1.89 | 0.16 | 1.10 | 0.46-2.64 | 0.82 |
| BMI >30 kg/m^2^ (Yes/No) | 1.07 | 0.80-1.45 | 0.62 | 1.01 | 0.75-1.37 | 0.92 |
| Country (Italy vs Greece) | 1.50 | 0.95-2.37 | 0.08 | 1.56 | 0.98-2.49 | 0.06 |

Abbreviations BMI: body mass index; CI: confidence internal

**Table S8 Changes of the Sequential Organ Failure Assessment score at day 7 from baseline**

Univariate and multivariate ordinal regression analyses are shown. Co-variates entered in the multivariate model were those used for stratified randomization according to the received advice by the COVID-ETF of the EMA. The analysis involves patients who remained hospitalized by day 7

| **Variable** | **Univariate analysis** | | | **Multivariate analysis** | | |
| --- | --- | --- | --- | --- | --- | --- |
|  | **Odds ratio** | **95% CIs** | **P-value** | **Odds ratio** | **95% CIs** | **P-value** |
| Group of treatment (Anakinra vs placebo) | 0.63 | 0.46-0.86 | 0.001 | 0.64 | 0.47-0.88 | 0.007 |
| Intake of dexamethasone (Yes/No) | 1.11 | 0.76-1.62 | 0.58 | 0.58 | 0.24-1.46 | 0.25 |
| Severe COVID-19 by WHO (Yes/No) | 1.25 | 0.86-1.81 | 0.25 | 1.89 | 0.76-4.65 | 0.17 |
| BMI >30 kg/m^2^ (Yes/No) | 1.25 | 0.92-1.69 | 0.15 | 1.26 | 0.92-1.71 | 0.14 |
| Country (Italy vs Greece) | 0.94 | 0.58-1.52 | 0.81 | 0.88 | 0.55-2.34 | 0.64 |

Abbreviations BMI: body mass index; CI: confidence internal

**Table S9. Complete list of serious treatment-emergent adverse events (TEAE) Classified by System**

|  | **SoC+ Placebo (n=189)** | **SoC+ Anakinra**  **(n=405)** | **P-value** |
| --- | --- | --- | --- |
| At least one serious TEAE, n (%) | 41 (21.2) | 68 (16.5) | 0.17 |
| Type of serious TEAE, n (%) |  |  |  |
| Infections and infestations, total | 25 (13.0) | 31 (7.5) | 0.035 |
| Ventilator-associated pneumonia | 14 (7.4) | 14 (3.5) | 0.039 |
| Related to the study drug | 2 (1.1) | 2 (0.5) |  |
| Bloodstream infection | 6 (3.2) | 12 (3.0) | >0.99 |
| Related to the study drug | 0 (0) | 0 (0) |  |
| *Clostridioides difficile* infection | 1 (0.5) | 1 (0.2) | 0.54 |
| Related to the study drug | 0 (0) | 0 (0) |  |
| Septic Shock and multiple organ dysfunction | 1 (0.5) | 1 (0.2) | 0.54 |
| Related to the study drug | 0 (0) | 0 (0) |  |
| Probable nosocomial infections | 4 (2.1) | 10 (2.5) | >0.99 |
| Related to the study drug | 1 (0.5) | 1 (0.2) |  |
| Acute pyelonephritis | 4 (2.1) | 5 (1.2) | 0.48 |
| Related to the study drug | 1 (0.5) | 1 (0.2) |  |
| Intrabdominal infection | 1 (0.5) | 2 (0.5) | >0.99 |
| Related to the study drug | 0 (0) | 0 (0) |  |
| Diagnosis of chronic hepatitis B | 0 (0) | 1 (0.2) | >0.99 |
| Related to the study drug | 0 (0) | 0 (0) |  |
| New hospital admissions | 1 (0.5) | 0 (0) | 0.32 |
| Related to the study drug | 0 (0) | 0 (0) |  |
| Acute kidney injury | 1 (0.5) | 4 (1.0) | >0.99 |
| Related to the study drug | 0 (0) | 1 (0.2) |  |
| Gastrointestinal hemorrhage | 1 (0.5) | 0 (0) | 0.32 |
| Related to the study drug | 0 (0) | 0 (0) |  |
| Anaphylactic shock | 0 (0) | 1 (0.2) | >0.99 |
| Related to the study drug | 0 (0) | 0 (0) |  |
| Lung, heart and vessels |  |  |  |
| Pulmonary embolism | 4 (2.1) | 7 (1.7) | 0.75 |
| Related to the study drug | 0 (0) | 0 (0) |  |
| Vascular thrombosis | 0 (0) | 1 (0.2) | >0.99 |
| Related to the study drug | 0 (0) | 0 (0) |  |
| Pneumomediastinum | 1 (0.5) | 3 (0.7) | >0.99 |
| Related to the study drug | 0 (0) | 0 (0) |  |
| Pneumothorax | 2 (1.1) | 1 (0.2) | 0.24 |
| Related to the study drug | 0 (0) | 0 (0) |  |
| Pulmonary fibrosis | 0 (0) | 1 (0.2) | >0.99 |
| Related to the study drug | 0 (0) | 0 (0) |  |
| Lung hemorrhage | 1 (0.5) | 0 (0) | 0.32 |
| Related to the study drug | 0 (0) | 0 (0) |  |
| Sinus bradycardia | 1 (0.5) | 2 (0.5) | >0.99 |
| Related to the study drug | 0 (0) | 0 (0) |  |
| Atrial fibrillation | 1 (0.5) | 2 (0.5) | 1.00 |
| Related to the study drug | 0 (0) | 0 (0) |  |
| Atrio-ventricular block | 1 (0.5) | 0 (0) | 0.32 |
| Related to the study drug | 0 (0) | 0 (0) |  |
| Ischemic stroke | 0 (0) | 2 (0.5) | >0.99 |
| Related to the study drug | 0 (0) | 0 (0) |  |
| Metabolic and electrolytes |  |  |  |
| Hyperglycemia | 2 (1.1) | 1 (0.2) | 0.24 |
| Related to the study drug | 0 (0) | 0 (0) |  |
| Hypoglycemia | 1 (0.5) | 2 (0.5) | >0.99 |
| Related to the study drug | 0 (0) | 0 (0) |  |
| Hypernatremia | 1 (0.5) | 4 (1.0) | >0.99 |
| Related to the study drug | 0 (0) | 0 (0) |  |
| Hyponatremia | 0 (0) | 2 (0.5) | >0.99 |
| Related to the study drug | 0 (0) | 0 (0) |  |
| Hyperkalemia | 1 (0.5) | 0 (0) | 0.32 |
| Related to the study drug | 0 (0) | 0 (0) |  |
| Hypocalcemia | 0 (0) | 1 (0.2) | >0.99 |
| Related to the study drug | 0 (0) | 0 (0) |  |
| Blood and lymphatic tissue |  |  |  |
| INR increase | 1 (0.5) | 1 (0.2) | 0.54 |
| Related to the study drug | 0 (0) | 0 (0) |  |
| Prolongation of aPTT | 2 (1.1) | 1 (0.5) | 0.24 |
| Related to the study drug | 0 (0) | 0 (0) |  |
| Decrease of fibrinogen | 1 (0.5) | 0 (0) | 0.32 |
| Related to the study drug | 0 (0) | 0 (0) |  |
| Increase of LFTs | 2 (1.1) | 4 (1.0) | >0.99 |
| Related to the study drug | 1 (0.5) | 2 (0.5) |  |
| Anemia | 3 (1.6) | 3 (0.7) | 0.39 |
| Related to the study drug | 0 (0) | 0 (0) |  |
| Neutropenia | 0 (0) | 1 (0.2) | >0.99 |
| Related to the study drug | 0 (0) | 1 (0.2) |  |
| Lymphopenia | 0 (0) | 3 (0.7) | 0.56 |
| Related to the study drug | 0 (0) | 1 (0.2) |  |

Abbreviations: aPTT: activated partial thromboplastin time; INR: international normalized ratio; LFTs: liver function tests; SoC: standard-of-care

**Table S10. Complete list of non-serious treatment-emergent adverse events (TEAE) Classified by System**

|  | **Standard-of-care + Placebo (N=189)** | **Standard-of-care + Anakinra**  **(N=405)** | **P Value** |
| --- | --- | --- | --- |
| At least an adverse event — no. (%) | 170 (90.4) | 352 (87.8) | 0.40 |
| Type of adverse event — no. (%) |  |  |  |
| Blood and lymphatic tissue |  |  |  |
| Leukopenia | 5 (2.6) | 14 (3.5) | 0.63 |
| Grade 1 | 4 (2.1) | 12 (3.0) | 0.60 |
| Grade 2 | 0 (0.0) | 1 (0.2) | >0.99 |
| Grade 3 | 1 (0.5) | 1 (0.2) | >0.99 |
| Neutropenia | 1 (0.5) | 12 (3.0) | 0.07 |
| Grade 1 | 1 (0.5) | 8 (2.0) | 0.28 |
| Grade 2 | 0 (0.0) | 4 (1.0) | 0.31 |
| Anemia | 37 (19.6) | 58 (14.3) | **<0.001** |
| Grade 1 | 32 (16.9) | 52 (12.8) | 0.21 |
| Grade 2 | 2 (1.1) | 6 (1.5) | >0.99 |
| Grade 3 | 3 (1.6) | 0 (0.0) | **0.03** |
| Thrombocytopenia | 4 (2.1) | 9 (2.2) | >0.99 |
| Grade 1 | 2 (1.1) | 6 (1.5) | >0.99 |
| Grade 2 | 1 (0.5) | 2 (0.5) | >0.99 |
| Grade 3 | 1 (0.5) | 1 (0.2) | >0.99 |
| Thrombocytosis | 13 (6.9) | 24 (5.9) | 0.72 |
| Grade 1 | 11 (5.8) | 24 (5.9) | >0.99 |
| Grade 2 | 2 (1.1) | 0 (0.0) | 0.10 |
| Skin and dermis |  |  |  |
| Reaction at injection site | 0 (0.0) | 2 (0.5) | 0.57 |
| Grade 1 | 0 (0.0) | 2 (0.5) | 0.57 |
| Rash | 3 (1.5) | 15 (3.7) | 0.20 |
| Grade 1 | 2 (1.1) | 11 (2.7) | 0.24 |
| Grade 2 | 1 (0.5) | 4 (1.0) | 0.68 |
| Gastrointestinal tract and liver |  |  |  |
| Nausea, vomiting | 1 (0.5) | 9 (2.2) | 0.18 |
| Grade 1 | 0 (0.0) | 8 (2.0) | 0.06 |
| Grade 2 | 1 (0.5) | 1 (0.2) | >0.99 |
| Constipation | 16 (8.5) | 39 (9.6) | 0.76 |
| Grade 1 | 15 (7.9) | 35 (8.6) | 0.87 |
| Grade 2 | 1 (0.5) | 2 (0.5) | >0.99 |
| Grade 3 | 0 (0.0) | 2 (0.5) | 0.57 |
| Diarrhea | 8 (4.2) | 14 (3.5) | 0.82 |
| Grade 1 | 7 (3.7) | 13 (3.2) | 0.81 |
| Grade 2 | 1 (0.5) | 1 (0.2) | >0.99 |
| Elevation of liver function tests | 63 (33.3) | 145 (35.8) | 0.58 |
| Grade 1 | 48 (25.4) | 111 (27.4) | 0.62 |
| Grade 2 | 11 (5.8) | 24 (5.9) | >0.99 |
| Grade 3 | 4 (2.1) | 10 (2.5) | >0.99 |
| Cardiovascular |  |  |  |
| Bradycardia | 19 (10.1) | 36 (8.9) | 0.76 |
| Grade 1 | 15 (7.9) | 31 (7.7) | >0.99 |
| Grade 2 | 3 (1.6) | 4 (1.0) | 0.69 |
| Grade 3 | 1 (0.5) | 1 (0.2) | >0.99 |
| Central nervous system |  |  |  |
| Headache | 8 (4.2) | 16 (4.0) | >0.99 |
| Grade 1 | 7 (3.7) | 13 (3.2) | 0.81 |
| Grade 2 | 1 (0.5) | 1 (0.2) | >0.99 |
| Grade 3 | 0 (0.0) | 1 (0.2) | >0.99 |
| Anxiety | 11 (5.8) | 33 (8.2) | 0.40 |
| Grade 1 | 8 (4.2) | 22 (5.4) | 0.56 |
| Grade 2 | 3 (1.6) | 11 (2.7) | 0.56 |
| Delirium | 2 (1.1) | 3 (0.7) | >0.99 |
| Grade 1 | 1 (0.5) | 0 (0.0) | 0.32 |
| Grade 2 | 0 (0.0) | 2 (0.5) | 0.57 |
| Grade 3 | 1 (0.5) | 1 (0.2) | >0.99 |
| Creatinine increase | 9 (4.8) | 17 (4.2) | 0.83 |
| Grade 1 | 4 (2.1) | 17 (4.2) | 0.24 |
| Grade 2 | 3 (1.6) | 0 (0.0) | **0.03** |
| Grade 3 | 2 (1.1) | 0 (0.0) | 0.10 |
| Metabolic and electrolytes |  |  |  |
| Hyperglycemia | 76 (40.2) | 148 (36.5) | 0.41 |
| Grade 1 | 61 (32.3) | 114 (28.1) | 0.33 |
| Grade 2 | 9 (4.8) | 19 (4.7) | >0.99 |
| Grade 3 | 6 (3.2) | 15 (3.7) | 0.82 |
| Hyponatremia | 23 (12.2) | 32 (7.9) | 0.13 |
| Grade 1 | 22 (11.6) | 28 (6.9) | 0.06 |
| Grade 2 | 1 (0.5) | 3 (0.7) | >0.99 |
| Grade 3 | 0 (0.0) | 1 (0.2) | >0.99 |
| Hypernatremia | 17 (9.0) | 46 (11.4) | 0.40 |
| Grade 1 | 14 (7.4) | 31 (7.7) | >0.99 |
| Grade 2 | 2 (1.1) | 9 (2.2) | 0.52 |
| Grade 3 | 1 (0.5) | 6 (1.5) | 0.44 |
| Hypokalemia | 12 (6.3) | 11 (2.7) | **0.04** |
| Grade 1 | 11 (5.8) | 9 (2.2) | **0.03** |
| Grade 2 | 1 (0.5) | 2 (0.5) | >0.99 |
| Hyperkalemia | 13 (6.9) | 36 (8.9) | 0.43 |
| Grade 1 | 7 (3.7) | 21 (5.2) | 0.54 |
| Grade 2 | 5 (2.6) | 10 (2.0) | >0.99 |
| Grade 3 | 1 (0.5) | 5 (1.2) | 0.67 |
| Hypercalcemia | 1 (0.5) | 4 (1.0) | 0.68 |
| Grade 1 | 1 (0.5) | 3 (0.6) | >0.99 |
| Grade 2 | 0 (0.0) | 1 (0.2) | >0.99 |
| Hypocalcemia | 20 (10.6) | 32 (7.9) | 0.35 |
| Grade 1 | 14 (7.4) | 19 (4.7) | 0.25 |
| Grade 2 | 6 (3.2) | 11 (2.7) | 0.79 |
| Grade 3 | 0 (0.0) | 2 (0.5) | 0.57 |
| Hypermagnesemia | 1 (0.5) | 2 (0.5) | >0.99 |
| Grade 1 | 1 (0.5) | 2 (0.5) | >0.99 |
| Hypomagnesemia | 1 (0.5) | 3 (0.7) | >0.99 |
| Grade 1 | 1 (0.5) | 3 (0.7) | >0.99 |
